# A Multitask Approach for Automated Detection and Segmentation of Thyroid Nodules in Ultrasound Images

**DOI:** 10.1101/2023.01.31.23285223

**Authors:** Ashwath Radhachandran, Adam Kinzel, Joseph Chen, Vivek Sant, Maitraya Patel, Rinat Masamed, Corey W. Arnold, William Speier

**Author notes:** This work has been submitted to the IEEE for possible publication. Copyright may be transferred without notice, after which this version may no longer be accessible.

## Abstract

An increase in the incidence and diagnosis of thyroid nodules and thyroid cancer underscores the need for a better approach to nodule detection and risk stratification in ultrasound (US) images that can reduce healthcare costs, patient discomfort, and unnecessary invasive procedures. However, variability in ultrasound technique and interpretation makes the diagnostic process partially subjective. Therefore, an automated approach that detects and segments nodules could improve performance on downstream tasks, such as risk stratification.Current deep learning architectures for segmentation are typically semi-automated because they are evaluated solely on images known to have nodules and do not assess ability to identify suspicious images. However, the proposed multitask approach both detects suspicious images and segments potential nodules; this allows for a clinically translatable model that aptly parallels the workflow for thyroid nodule assessment. The multitask approach is centered on an anomaly detection (AD) module that can be integrated with any U-Net architecture variant to improve image-level nodule detection. Ultrasound studies were acquired from 280 patients at UCLA Health, totaling 9,888 images, and annotated by collaborating radiologists. Of the evaluated models, a multi-scale UNet (MSUNet) with AD achieved the highest F1 score of 0.829 and image-wide Dice similarity coefficient of 0.782 on our hold-out test set. Furthermore, models were evaluated on two external validations datasets to demonstrate generalizability and robustness to data variability. Ultimately, the proposed architecture is an automated multitask method that expands on previous methods by successfully both detecting and segmenting nodules in ultrasound.

## I. Introduction

There has been a recent rise in thyroid cancer incidence, making it the fifth most common malignancy amongst women in the United States between 2015-2019. [1] By age 50, 50–60% of the population have one or more thyroid nodules incidentally discovered on imaging performed for an unrelated indication. [2] Although 90% of these detected nodules are actually clinically insignificant and benign, uncertainty in radiologic evaluation can lead to unnecessary biopsies. [3] Ultrasound (US) imaging is the primary imaging modality used to assess the thyroid along with the morphology and internal architecture of any present nodules. [4] When evaluating nodules, the first step involves detecting if and where a nodule is present on a series of US images. However, the presence of various technical limitations and artifacts in US images such as vague nodule boundaries, poor contrast, variable probe positioning, and non-standardized spatial resolution, makes the diagnostic process partially subjective. These limitations lead to high inter-radiologist variability [5] in risk stratification, and an increased need for invasive diagnostic procedures such as fine needle aspiration biopsies (FNABs). [6] And oftentimes, FNABs can be inconclusive, leading to unnecessary healthcare costs and complications for patients. [7]

In recent years, an increase in the development of computeraided diagnosis (CAD) methods using deep learning has proven useful for image analysis tasks in biomedicine. [8]–[10] Deep learning can be especially useful in eliminating variability and enhancing diagnostic reliability in thyroid cancer risk assessment by improving nodule localization techniques. [11] Multiple groups have explored different deep learning architectures to address localization through the segmentation of thyroid nodules. [12]–[15] While these existing models demonstrate promising results, they are semi-automated because they are evaluated solely on images known to have nodules and do not assess ability to identify suspicious images. Thus, most current work tackles a more narrow task, which despite performing well, does not translate in terms of clinical applicability. For such a segmentation model to be implemented successfully, it would require a radiologist to first identify suspicious images, on which the model would make nodule segmentation predictions. However, in the clinical setting, when a radiologist evaluates a set of US images and identifies an image with a potential nodule, they are simultaneously deciding where the nodule is located. Without incorporating a method for image-wide detection, a model that purely segments is unlikely to have a tangible clinical impact.

As a standalone algorithm, image-wide detection of thyroid nodules on ultrasound has not been extensively studied. Moreover, such an algorithm would require a secondary purpose such as nodule localization to augment the clinical workflow. [16] In order to improve detection of anomalous images and elevate the clinical utility of a segmentation model, this work introduces a multitask approach through an anomaly detection (AD) module to be used in conjunction with any U-Net architecture variant. Previously, Jain et al. [17] proposed a two-stage model that performed AD to filter out non-suspicious capsule endoscopy images and then segmented the remaining images. In a different study, Chen et al. [18] integrated a mask classification component into their dermoscopic image segmentation network to determine if a lesion was present. In that work, the mask classification output was only used to improve the training of model weights. The proposed AD module expands on this by not only making a binary classification of nodule presence, but multiplying the AD output with the nodule segmentation to post-process the prediction. By concurrently determining which images may have nodules and segmenting suspicious regions, our multitask approach allows for a clinically translatable model that aptly parallels the workflow for thyroid nodule assessment.

The AD module is integrated with a variety of state-of-the-art segmentation architectures to understand its impact on performance. A majority of past work has studied the UNet architecture, a gold standard for segmentation tasks. [8] However, the UNet implements the same convolutional filter size, resulting in a fixed receptive field which hampers the segmentation of objects that vary in size. In response to this issue, Su et al. proposed the multi-scale UNet (MSUNet) [19], which introduces a multi-scale block in each layer of the encoder to fuse the outputs of convolution kernels with different receptive fields. The multi-scale block helps capture more diverse features and detailed spatial information from the input. This innovation is especially relevant for thyroid nodule segmentation since the target presents in diverse shapes and sizes. MSUNet was shown to demonstrate consistent performance when evaluated on various medical image segmentation datasets, one of which was a collection of breast ultrasound lesion images. However, the architecture was not specifically evaluated on ultrasound images for thyroid nodule segmentation. This paper extends the application of MSUNet and introduces the AD module to create a more clinically impactful segmentation method. In summary, the main contributions of this work are as follows:

1. To more closely parallel the distribution of images seen in medical practice and gather a more clinically relevant metric, segmentation performance is measured on images with and without nodules. In contrast to previous research, negative images (those without nodules) are included during evaluation.
2. The AD module is integrated with the segmentation model architecture to assist in image-wide nodule detection. Thus, the proposed multitask method steps towards an automated segmentation method that does not rely on guidance from radiologists.
3. MSUNet, with and without the AD module, is compared against various state-of-the-art models for nodule segmentation.
4. The evaluated architectures are trained and tested on a novel thyroid ultrasound dataset. Furthermore, they are externally validated on two publicly available thyroid ultrasound datasets, demonstrating model stability and robustness to data variability.

## II. METHODS

### A. Dataset Descriptions

#### 1) UCLA Dataset

UCLA Health has implemented a standardized protocol for the acquisition of thyroid US images. Over the past three years, patients who have had an US examination following this protocol have had their imaging collected and aggregated in the UCLA Thyroid RadPath research dataset. This study consists of 280 patients from this dataset, where each patient has a set of 20-40 US images that span a variety of anatomic regions and ultrasound probe orientations. Each US image has been manually segmented by two radiologists and validated by a more senior radiologist at UCLA.

#### 2) Digital Database Thyroid Image (DDTI)

DDTI is a public, open access dataset from the IDIME Ultrasound Department, one of Colombia’s largest diagnostic imaging centers. [20] This dataset contains 480 images from 290 patients. Each of these images is accompanied by a nodule annotation made by a radiology resident. Preprocessing of the original 480 images resulted in 610 images (images that contained two frames in one were split and considered separately).

#### 3) Stanford CINE

The Stanford CINE dataset contains 192 cine clips from 167 patients with biopsy-confirmed thyroid nodules. [21] The cine clips in this dataset included radiologist-annotated segmentations and captured a single nodule. On average, each cine clip was composed of 90 frames, but the total number of frames was downsampled by a factor of 10 due to redundancy in image content. The frames from the cine clips were extracted and treated as independent images.

### B. AD Module

The central innovation of this approach is the integration of the AD module with any variant of the U-Net architecture, such as MSUNet, to enable a multitask model for thyroid nodule segmentation on ultrasound images (Fig. 2). The contracting path of a U-Net based architecture is engineered to extract a high-level representation of an input image. The AD module flattens this feature map and passes it through a four-layer fully connected network (FCN) that performs a binary classification of nodule presence. The FCN includes Dropout layers, to prevent overfitting and LeakyReLU activation, to maintain a non-zero gradient during training. Then, the FCN output is rescaled between 0 and 1 using a sigmoid function. The AD module leverages the feature rich image encoding, without requiring unnecessary feature engineering or processing steps, to make a streamlined classification of nodule presence with minimal overhead. The expansive path of the U-Net architecture also takes the image encoding from the contracting path, but reconstructs a predicted segmentation mask. Thus, for every input image, MSUNet with the AD module (MSUNet-AD) will predict a nodule mask and AD output. During evaluation, an empirically determined threshold (details under Sensitivity Analysis in Results) is used to binarize the AD output, which corresponds to MSUNet-AD’s prediction of whether or not the input image contains a nodule. The AD output is then multiplied with the binary mask to either maintain the segmentation prediction or convert it into an empty mask.

**Fig. 1.**
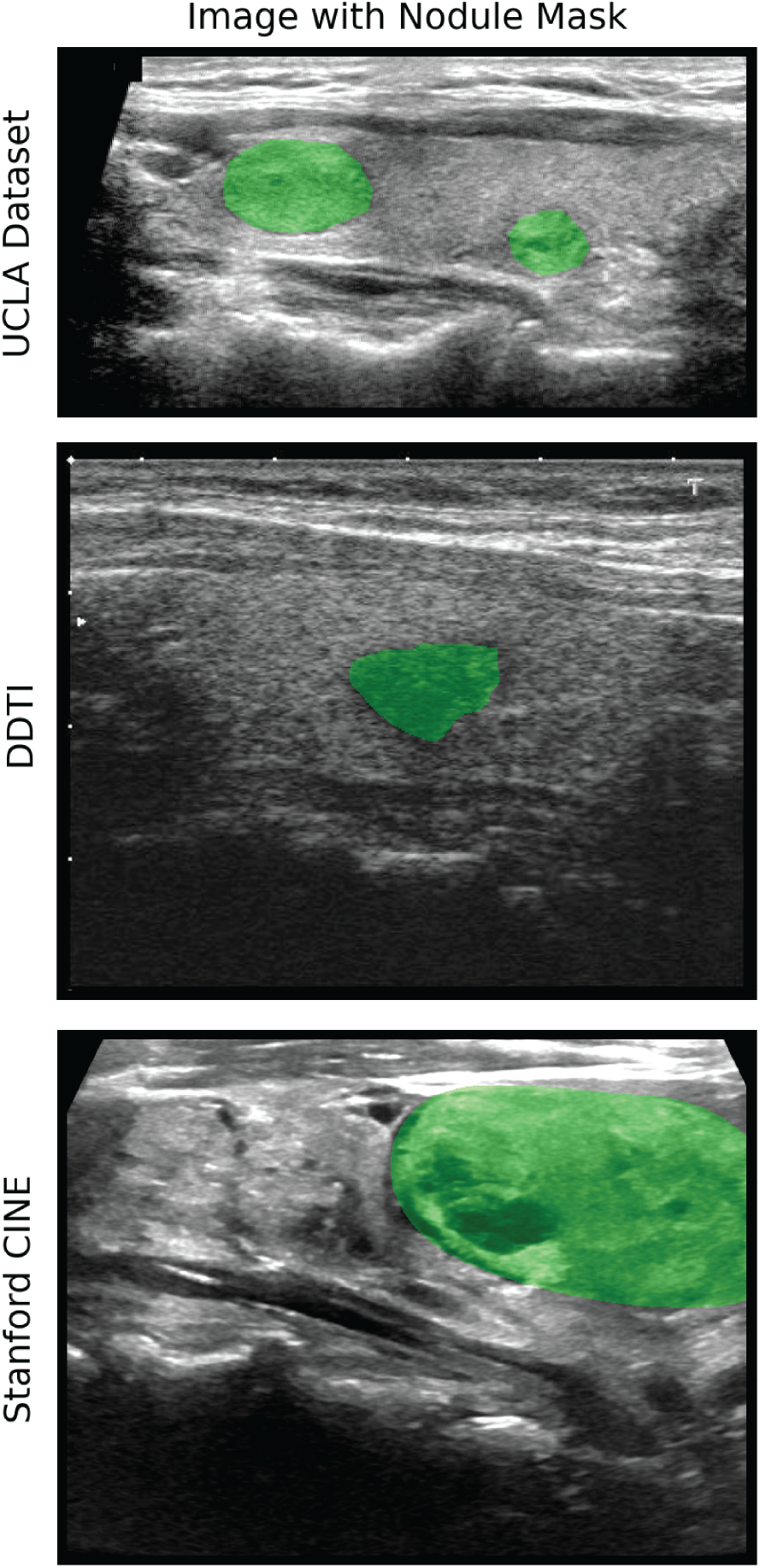
Example of an image with corresponding annotation mask from each dataset.

**Fig. 2.**
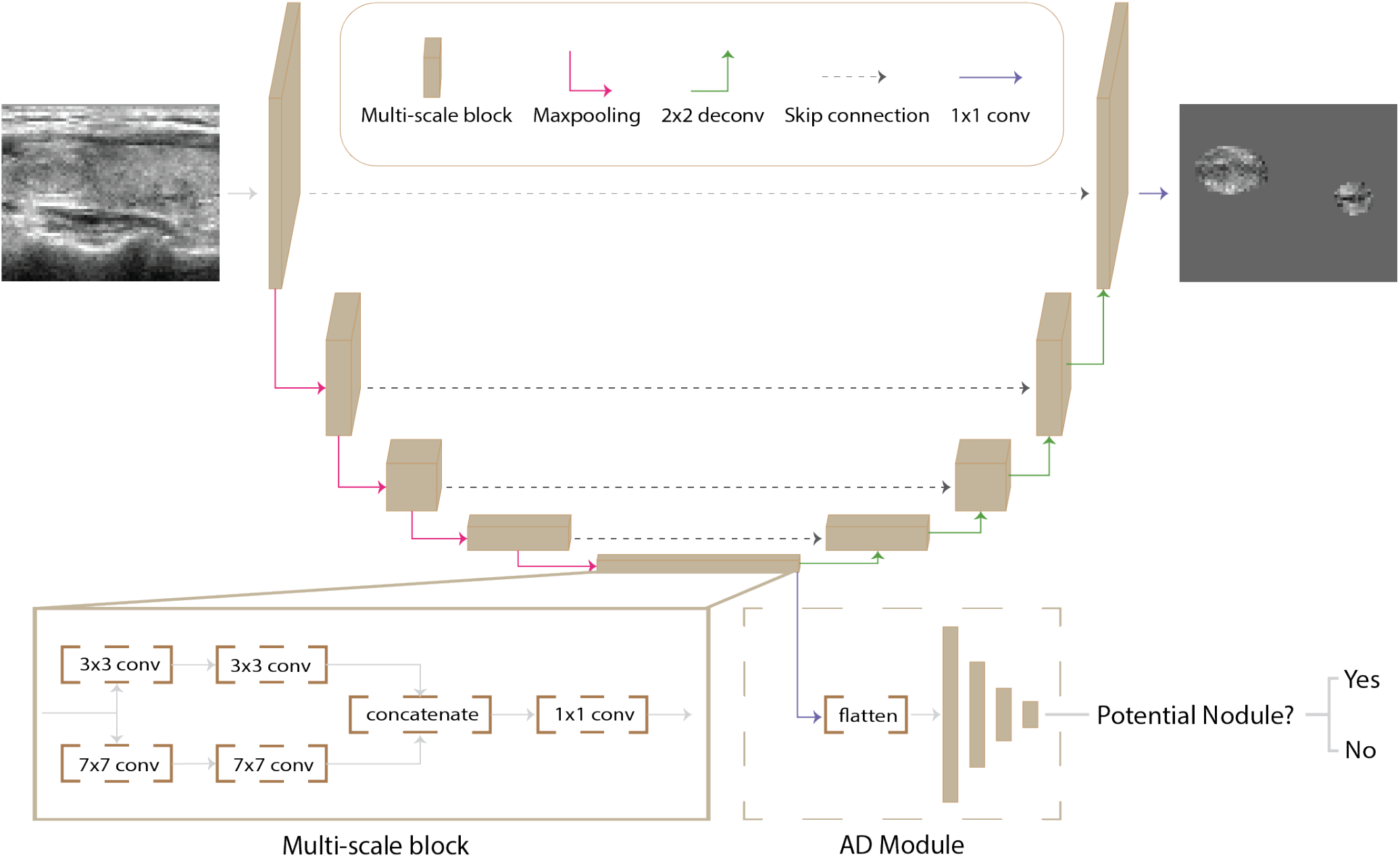
An overview of the model architecture for MSUNet-AD, with an in-depth look at the operations in each multi-scale block (bottom left) and in the AD module (bottom right).

### C. MSUNet

The UNet has been a standard architecture used for medical image segmentation. The encoding pathway consists of multiple layers, each of which contain a sequence (block) of 3×3 convolutions that extract relevant image features. However, using the same 3×3 kernel size for encoding pathway convolutions will lead to a fixed receptive field size, which can cause shortcomings in feature extraction during the encoding pathway and feature recovery during the decoding pathway. Small receptive fields typically excel at segmenting small objects, but struggle at localization, whereas large receptive fields are more conducive to segmenting and localizing larger objects. [22] In response to this tradeoff, Su et al. introduced MSUNet and the idea of the multi-scale block, which incorporates convolution sequences with different kernel sizes, allowing for the extraction of more diverse features. The multi-scale block consists of parallel paths each applying a double convolution, one with kernel size 3×3 and one with kernel size 7×7. The outputs of these layers are concatenated, a 1×1 convolution is applied and the resulting matrix is fed into the next layer.

### D. Segmentation Architectures

#### 1) Semantic Guided UNet (SGUNet)

SGUNet [14] is a UNet inspired architecture designed specifically for thyroid nodule segmentation in ultrasound. Their primary innovation is the SGM module, which reduces noise interference inherent to ultrasound imaging that may be propagated in the encoding layer convolutions as well as in the skip connections to the decoder layers. A SGM module is included after each decoding layer to convert the upsampled decoding into a one channel feature map. This one channel map is then concatenated with the previous decoding layer’s SGM module output. A 1×1 convolution is applied to the resulting two channel map to result in that decoding layer’s one channel semantic map. SGUNet was re-implemented in PyTorch, trained on the UCLA dataset and used as a baseline to compare against the proposed architecture.

#### 2) MICCAI 2020 TN-SCUI Challenge Winner (Wang et al.)

The Wang et al. model [23] is a two stage cascaded framework that was developed for single-target segmentation of thyroid nodules. The stage 1 network is trained to predict a nodule given an ultrasound image, while the stage 2 network is trained on an image crop containing the region of interest derived from the ground truth annotation. Thus, during evaluation the two stages can be used sequentially, where stage 1 makes an initial prediction to localize the nodule, and stage 2 performs a finer segmentation of the identified nodule. Pretrained weights for stage 1 and stage 2 from training on the TN-SCUI challenge dataset were used to compare against Wang et al. when trained and evaluated on the UCLA dataset. Lastly, the proposed AD module could not be integrated with the Wang et al. model. Instead, a similar AD module was trained in parallel and during evaluation, its binary output was used to post-process the predicted mask. The AD output is multiplied with the mask to either maintain the prediction or convert it to an empty mask.

#### 3) Additional Baselines

The proposed architecture was compared against standard baselines that are commonly considered in segmentation papers. These additional baselines include the UNet, [8] UNet++ (Nested UNet), [24] UNet with pretrained ResNet50 encoder (SResUNet) and Attention UNet (AttUNet). [25]

### E. Model Training

An 80:20 train-test split was applied at the patient level to the UCLA dataset. 10% of the training set patients made up the validation set. Images with certain artifacts, such as transducer information, vascular flow assessments (Doppler images) or caliper markings, were removed. Each of the remaining images went through removal of protected health information, contrast enhancement and whitening. Image counts after applying this preprocessing pipeline are presented in Table I.

**TABLE I.** Patient and image counts for train, validation and test splits. Images with at least one ground truth nodule are referred to as positive images. The images without nodules are negative images. The percentage of positive images are included in the last column.

| Split | Patients | Images | Positive Images |
| --- | --- | --- | --- |
| train | 202 | 7097 | 2892 (40.7%) |
| validation | 22 | 793 | 332 (41.9%) |
| test | 56 | 1998 | 766 (38.3%) |

Image inputs for model training, validation, and testing were resized to 256 × 256. Image augmentation was performed through random flipping, rotation, and addition of Gaussian noise. The Adam optimizer was used with an initial learning rate of 0.0005 and batch size of 64. The learning rate was decreased by a factor of 10 if the validation loss did not improve for 5 epochs. The segmentation models without AD were trained using DiceBCE loss. A value of 0.5 is empirically assigned to w so that both the Dice loss and pixel-level BCE loss (P-BCE) have equal weighting. Upon integration of the AD module with the segmentation model, BCE loss was used for the AD classification task (AD-BCE). Equation (1) shows how these losses are used in concordance to train the multitask model. All models were developed using PyTorch 1.11.0 and trained on a NVIDIA DGX-1.

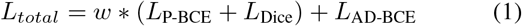

### F. Evaluation Metrics

Various evaluation metrics were used to quantify segmentation performance. The image-wide metrics are are Dice similarity coefficient (*DSC*_*all*_), precision, recall and F1 score. Precision, recall, and F1 score are calculated using a slightly different definition for true positives (non-empty prediction that overlaps with ground truth positive), true negatives (empty prediction for a ground truth negative), false positives (non-empty prediction for a ground truth negative) and false negatives (empty prediction for a ground truth positive or non-empty prediction that does not overlap with ground truth positive). *DSC*_*all*_ is the average Dice similarity coefficient (DSC) across all evaluated images. Because the Dice coefficient is undefined in cases where no nodule is present, Laplace smoothing (*λ*=1e-6) is applied to allow for calculation across all images. Thus, when DSC is calculated for a negative image, an empty predicted mask will lead to a DSC of 1, indicating a true negative. The positive image-level metrics are *DSC*_+_ and Intersection over Union (IoU). These metrics are averaged across only the positive images in the test set. When calculating DSC, as appears in (2), true positives (TP), false positives (FP) and false negatives (FN) are determined using the traditional pixel-level definition.

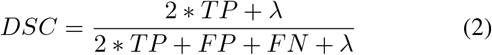

The Wilcoxon Rank Sum test was used to compare model performance based on DSC. Since the test set was determined based on patient-level split, there could be overlap in anatomical regions captured by a patient’s US images, which could unintentionally inflate the statistical power of the results. To address this shortcoming, the mean DSC for each patient’s positive images were calculated and the test statistic was calculated using these patient-level DSC averages instead of *DSC*_*all*_ or *DSC*_+_. For the Stanford CINE dataset, since adjacent frames tended to capture related anatomical views, DSC was averaged across the frames for each cine clip. For DDTI, although some patients had multiple images, they typically were of varying anatomical views so result aggregation was unnecessary.

## III. Results

### A. Segmentation Performance

The models were evaluated on the holdout test set with and without the AD module integrated into their architecture. MSUNet-AD had the highest *DSC*_*all*_ of 0.782 and F1 of 0.829 when compared against the other models with AD. In terms of *DSC*_+_, the models with AD performed between 0.571 and 0.627. In order to assess the effect of the AD module, further evaluation was performed on all the models without the AD module (Table II). When AD is integrated into the model architecture, there are increases in *DSC*_*all*_ and F1. For example, for MSUNet there was an increase in F1 from 0.683 to 0.829 and in *DSC*_*all*_ from 0.581 to 0.782. MSUNet also had a *DSC*_+_ of 0.726, while the other networks ranged between 0.621 and 0.711. Another observed trend when AD was integrated into the model architectures was a drop in *DSC*_+_ from values between 0.621 and 0.757 to values between 0.571 and 0.627 (*P<*0.05). The drop in performance on *DSC*_+_ can be attributed to variance in performance across different nodule sizes. Nodule size was determined by calculating the number of pixels a nodule(s) occupied on the image. The nodule sizes for all the positive images in the test set were aggregated and split into quartiles, where Q1 (smallest quartile) represents the smaller nodules and Q4 (largest quartile) represents the larger nodules. These quartiles are used to stratify the positive images in the test set and perform a more thorough evaluation.

**TABLE II.** Performance of models with and without AD on hold-out test set. Bolded values highlight the model with the best performance on a certain metric.

| Model Name | Image-Wide |  |  |  | Average Across Positive Images |  |
| --- | --- | --- | --- | --- | --- | --- |
| | $DSC_{all}$ | Precision | Recall | F1 | $DSC_+$ | IoU |
| UNet-AD | 0.775±0.357 | 0.807 | 0.822 | 0.814 | 0.609±0.340 | 0.514±0.316 |
| NestedUNet-AD | 0.762±0.364 | 0.795 | **0.850** | 0.821 | 0.597±0.335 | 0.498±0.308 |
| SResUNet-AD | 0.754±0.362 | 0.793 | 0.832 | 0.812 | 0.575±0.321 | 0.469±0.290 |
| Wang et al.-AD | 0.755±0.385 | 0.755 | 0.768 | 0.761 | 0.609±0.379 | 0.533±0.355 |
| SGUNet-AD | 0.722±0.390 | 0.731 | 0.811 | 0.769 | 0.571±0.344 | 0.474±0.313 |
| AttUNet-AD | 0.776±0.355 | 0.804 | 0.846 | 0.824 | 0.623±0.334 | 0.527±0.313 |
| MSU-Net-AD | **0.782±0.354** | **0.812** | 0.847 | **0.829** | **0.627±0.343** | **0.537±0.323** |
| UNet | 0.544±0.445 | 0.512 | 0.946 | 0.665 | 0.711±0.276 | 0.610±0.277 |
| NestedUNet | 0.539±0.444 | 0.508 | 0.944 | 0.661 | 0.710±0.271 | 0.606±0.271 |
| SResUNet | 0.259±0.369 | 0.369 | 0.941 | 0.530 | 0.673±0.277 | 0.563±0.271 |
| Wang et al. | 0.299±0.402 | 0.377 | **0.961** | 0.541 | **0.757±0.248** | **0.660±0.258** |
| SGUNet | 0.442±0.435 | 0.458 | 0.920 | 0.611 | 0.621±0.289 | 0.506±0.273 |
| AttUNet | 0.547±0.444 | 0.515 | 0.945 | 0.667 | 0.707±0.277 | 0.606±0.279 |
| MSU-Net | **0.581±0.442** | **0.535** | 0.944 | **0.683** | 0.726±0.271 | 0.628±0.273 |

A closer look at performance across the quartiles, upon addition of AD, shows that there is a larger drop in DSC on the smallest quartile than on the largest quartile. There were significant differences in DSC when AD was added between the majority of models on Q1 and Q2 (Fig. 3). One such case is the significant drop in DSC from MSUNet to MSUNet-AD in Q1 (*P*=0.001) and Q2 (*P*=0.002). An example of this can be seen in the 2nd row of Fig. 6, where MSUNet-AD and AttUNet-AD are unable to segment a smaller nodule, while their non-AD counterparts are at least able to localize it. However, the drops in DSC from MSUNet to MSUNet-AD were insignificant for Q3 (*P*=0.059) and Q4 (*P*=0.10). The more consistent performance on larger nodules can be seen in Fig. 6 (4th row), where all three models with and without AD are able to successfully segment the more prominent nodule. Ultimately, when AD is added to the other baseline models, the pattern of DSC falling more on the smallest quartile compared to the largest quartile is also observed. Moreover, there is less variability in DSC when considering performance on the largest quartile compared to the smallest quartile. An example of the greater variability in the smallest quartile can be seen in Fig. 6 (2nd row, 3rd row). In the 3rd row, MSUNet-AD is able to successfully segment a small nodule, whereas it struggles to segment a similarly sized nodule in the 2nd row example. Although the addition of the AD module leads to an increase in F1, it results in a drop in DSC that can be attributed to poorer segmentation of smaller nodules.

**Fig. 3.**
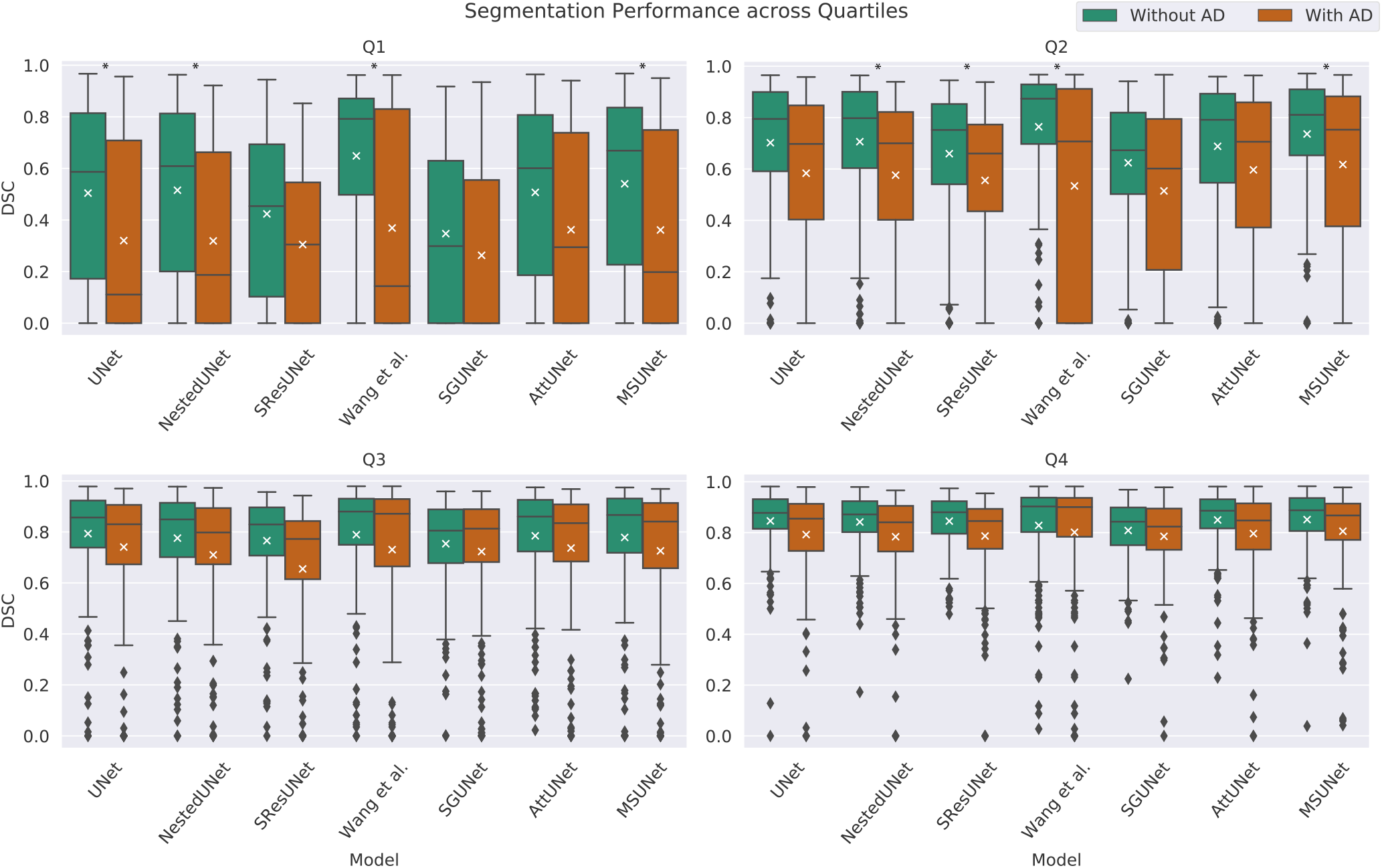
Box plot comparing segmentation performance by nodule size quartile between models with and without AD module. The plot represents the distribution of the DSC values (used to calculate ***DSC***_**+**_) for each quartile. A black asterisk above a comparison represents a statistically significant difference between models with and without the AD module. The white crosshair marks the mean of the distribution.

**Fig. 4.**
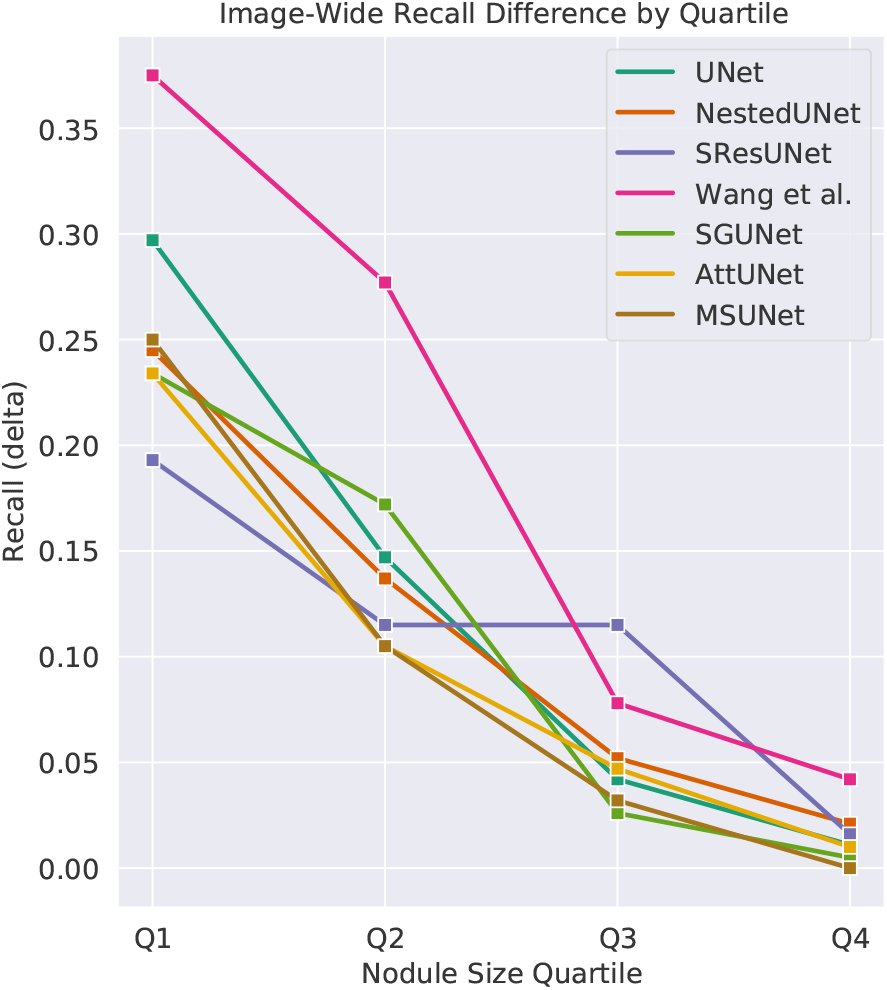
Image-wide recall was calculated for each model for each of the nodule size quartiles. The difference in recall between a model without and with AD is plotted for each quartile.

**Fig. 5.**
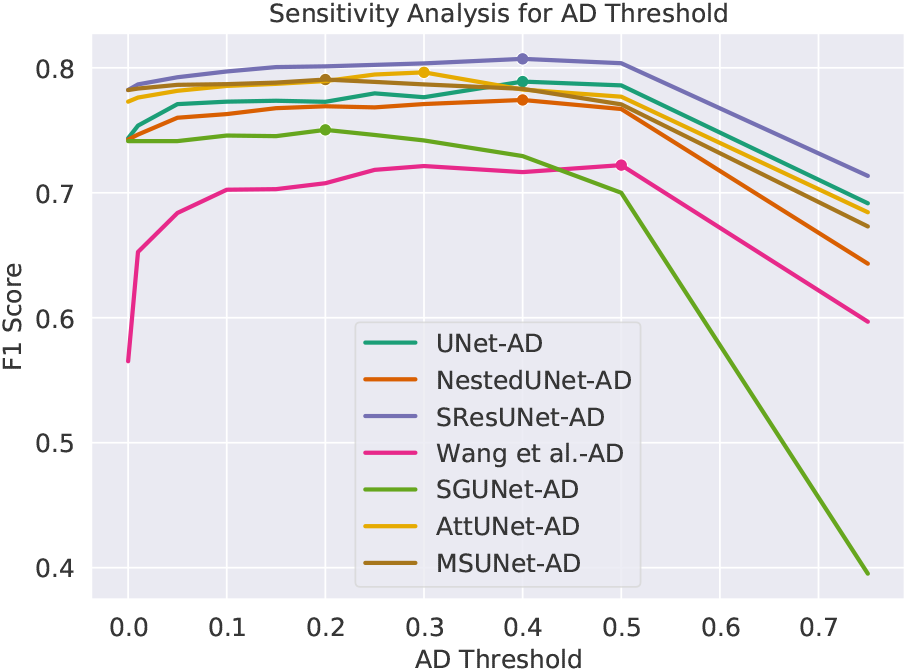
F1 performance across AD thresholds for the six models with AD. Dot marker represents the optimal threshold, where the best validation set F1 score is achieved for a given model.

**Fig. 6.**
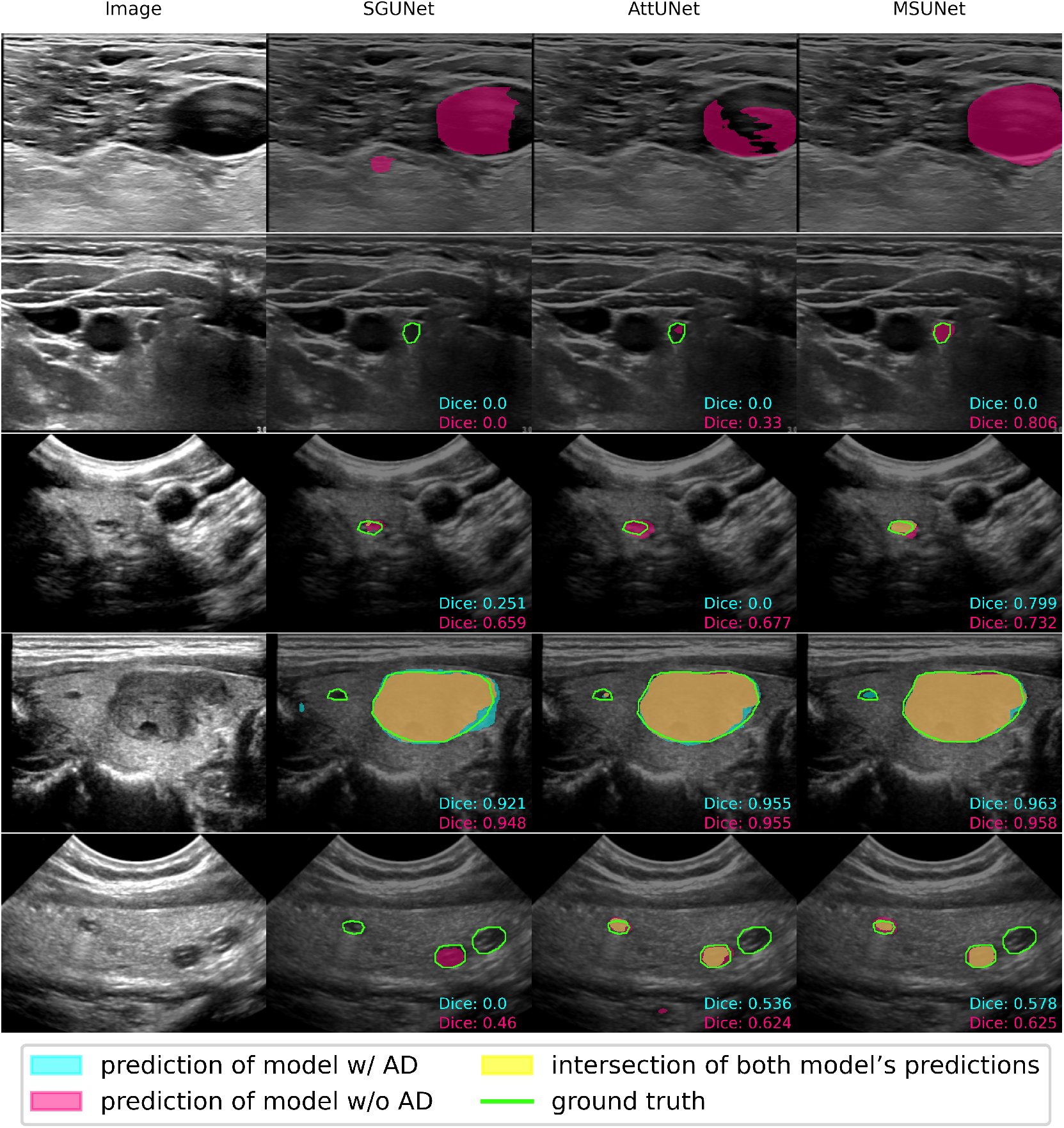
Examples of the difference in segmentation performance between three of the models evaluated. First row: All models without the AD module conflate a jugular vein as a nodule leading to a false positive. When the AD module is added, none of the models contain a prediction since the AD output correctly eliminates any predicted segmentations. Second row: An example of all models except MSUNet struggling to segment a small nodule. Third row: An example of a nodule where only MSUNet and MSUNet-AD exhibit agreement, whereas SGUNet-AD and AttUNet-AD struggle in comparison to their non-AD counterparts. Fourth row: An example of all models successfully segmenting the larger of two nodules. MSUNet-AD, AttUNet and AttUNet-AD are the only ones capable of detecting the smaller, adjacent nodule, but they struggle to completely segment it. Fifth row: A relatively difficult example with three different small nodules. All six models consistently miss the far right nodule. Here, AttUNet-AD and MSUNet-AD perform at par with the models without AD.

#### 1) Wang et al. Evaluation Results

The Wang et al. architecture was trained on the TN-SCUI dataset and evaluation on the TN-SCUI test set showed an increase in DSC when the stage 2 network was used with stage 1. Wang et al. made the weights of their trained model publicly available; when evaluated on the UCLA dataset, their model achieved a *DSC*_+_ of 0.708. When trained and tested on the UCLA dataset, there was a decrease in *DSC*_+_ from 0.757 to 0.635 when stage 1 and stage 2 were used in conjunction as opposed to just stage 1. Thus, the results for the Wang et al. model (Table II) are from the best performing permutation of the model architecture, which is when only stage 1 was used. Of the models without AD, the Wang et al. model had one of the lowest F1 scores of 0.541 and *DSC*_*all*_ of 0.299, but the highest *DSC*_+_ of 0.757. When AD was incorporated, the Wang et al. model had a *DSC*_*all*_ of 0.755, but the lowest F1 score of 0.761 and a drop in *DSC*_+_ to 0.609.

### B. Sensitivity Analysis

A sensitivity analysis was conducted to determine the best threshold for binarizing AD output. Various thresholds between 0 and 0.75 in increments of 0.05 were tested. Each network with AD was evaluated on the validation set and image-wide F1 scores were recorded using each of these different thresholds (Fig. 5). All the models with the AD module had an optimal AD threshold of either 0.2, 0.3, or 0.4, where they demonstrated best F1 performance on the validation set. There is a consistent plateau in F1 scores up until 0.5 for all models except SGUNet-AD, indicating that performance would not differ too drastically for most AD thresholds in that range.

### C. External Validation

The models with and without the AD module were evaluated on two external validation (EV) datasets, DDTI and Stanford CINE dataset. One difference between the UCLA dataset and these two datasets is that the latter does not have any negative images. Since the EV datasets only contain positive images, AD does not necessarily need to be performed. By setting the threshold at which the AD module output is binarized to 0, all images will be suspected to have a nodule(s) and it becomes possible to assess the segmentation ability of the models with AD. Unlike in the other models, the AD component for the Wang et al. architecture was trained independently. Thus when the threshold is 0, it eliminates the effect of the AD module. Hence, the EV results for the Wang et al. model with and without AD are equivalent.

The segmentation models without AD had a higher *DSC*_+_ compared to those with AD. For example, MSUNet had a *DSC*_+_ of 0.705 and 0.658, on DDTI and Stanford CINE respectively, whereas MSUNet-AD had a *DSC*_+_ of 0.527 and 0.569. For DDTI, the other models without AD had *DSC*_+_’s ranging between 0.587 and 0.728, while those with AD ranged between 0.494 and 0.576. For Stanford CINE, the other models without AD had *DSC*_+_’s ranging between 0.563 and 0.701, while those with AD ranged between 0.456 and 0.569 (Table III). *DSC*_+_ for MSUNet on DDTI was higher than that for AttUNet (*P<*0.001) and UNet (*P<*0.001). This was also the case for *DSC*_+_ for MSUNet on Stanford CINE compared to AttUNet (*P*=0.002) and UNet (*P*=0.006). *DSC*_+_ for MSUNet on the UCLA dataset was 0.726, which was higher than *DSC*_+_ of 0.705 (*P*=0.67) on DDTI and 0.658 on Stanford CINE (*P<*0.001). On the UCLA dataset, MSUNet-AD had a *DSC*_+_ of 0.627 which was higher than the *DSC*_+_ of 0.527 on DDTI (*P*=0.20) and 0.569 on Stanford CINE (*P*=0.016).

**TABLE 3.** Performance of models on DDTI and Stanford CINE datasets. Bolded values highlight the model with the best performance on a certain metric.

| Model Name | DDTI |  | Stanford CINE |  |
| --- | --- | --- | --- | --- |
|  | Average Across Positive Images |  | Average Across Positive Images |  |
| | $DSC_+$ | IoU | $DSC_+$ | IoU |
| UNet-AD | 0.494±0.242 | 0.364±0.223 | 0.456±0.264 | 0.335±0.236 |
| NestedUNet-AD | 0.502±0.247 | 0.372±0.227 | 0.476±0.275 | 0.356±0.244 |
| SResUNet-AD | **0.576±0.224** | **0.438±0.212** | 0.535±0.251 | 0.404±0.230 |
| SGUNet-AD | 0.513±0.257 | 0.384±0.232 | 0.530±0.271 | 0.405±0.248 |
| AttUNet-AD | 0.504±0.234 | 0.370±0.215 | 0.462±0.264 | 0.339±0.232 |
| MSU-Net-AD | 0.527±0.325 | 0.421±0.290 | **0.569±0.322** | **0.463±0.294** |
| UNet | 0.639±0.203 | 0.501±0.211 | 0.616±0.245 | 0.488±0.242 |
| NestedUNet | 0.614±0.195 | 0.470±0.195 | 0.609±0.258 | 0.483±0.249 |
| SResUNet | 0.604±0.214 | 0.466±0.215 | 0.601±0.251 | 0.472±0.242 |
| Wang et al. | **0.728±0.241** | **0.617±0.243** | **0.701±0.303** | **0.609±0.299** |
| SGUNet | 0.587±0.202 | 0.443±0.197 | 0.563±0.253 | 0.433±0.234 |
| AttUNet | 0.649±0.191 | 0.509±0.204 | 0.612±0.244 | 0.483±0.240 |
| MSU-Net | 0.705±0.191 | 0.575±0.210 | 0.658±0.269 | 0.542±0.267 |

## IV. Discussion and Conclusions

The AD module improved F1 score and *DSC*_*all*_ across the board when integrated with the segmentation models, with MSUNet-AD performing best in these two metrics. This increase can be attributed to the decrease in false positives at the image-wide level. Prior to the addition of AD, the Wang et al. model performed the best in terms of *DSC*_+_; however, the low *DSC*_*all*_ and precision, also seen in the other architectures without AD, demonstrated that the model’s predictions contained several false positives. Although AD helped increase precision, there was also an increase in false negatives, hence the decrease in recall and *DSC*_+_ for the models with AD. Further investigation into this pattern of decreasing *DSC*_+_ revealed that this drop was mainly due to an inability to segment smaller nodules (Fig. 3). This was seen across most models, as exemplified in both the quantitative disparities between Q1 and the other quartiles, and the segmentation examples (2nd row, Fig. 6). Moreover, the false negatives occurred with greater frequency in the smaller quartiles, as evidenced by the greater delta in image-wide recall between a model without and with AD in Q1 compared to in Q4 (Fig. 4). However, smaller nodules are less likely to be clinically significant [26], thus reducing the clinical consequences of the increase in false negatives due to AD. One limitation in these experiments was that the formula used to calculate size was pixel-based and not necessarily geared towards capturing the clinical definition of nodule size.

In regards to the EV experiments, there were notable advantages and disadvantages to the datasets used. DDTI was a favorable dataset because it contained multi-nodule images, which is comparable to real-world data and it followed an image collection protocol different from UCLA’s protocol, demonstrating the model’s independence from a specific image collection method. The main drawback of the Stanford CINE dataset was that it only contained clips of single nodules, which does not reflect real-world frequency. As detailed in the results, MSUNet-AD also outperformed the majority of other baselines on the EV datasets. It displayed similar *DSC*_+_ performance on the DDTI dataset, and despite falling short on the Stanford CINE dataset, the results indicated there was no severe overfitting to patterns found within images collected using UCLA’s standard US imaging protocol. Although the clinical implications of such models with AD need to be evaluated to understand how they would augment practice, these results indicate that MSUNet-AD is a versatile model that could have clinical applicability.

Through a multitask approach, MSUNet-AD advances current thyroid nodule segmentation methods with an automated technique that is designed to parallel clinical practice, and in doing so augment a radiologist’s workflow. The experimental results demonstrate that a previously developed architecture, MSUNet, upon the addition of AD, performs best in terms of image-wide detection and pixel-level nodule segmentation. Furthermore, the architectures with AD were evaluated on two external validation sets, DDTI and Stanford CINE, to demonstrate model stability and robustness to data variability. This technique can be extended to have further clinical application by extracting nodule features from segmented regions to develop a network that can perform an improved TI-RADS evaluation or automated biopsy prediction. Future work will aim to integrate the proposed multitask model into an end-to-end network that is also capable of nodule malignancy classification and further risk stratification. With rigorous evaluation, it would be possible to reduce the subjectivity in radiological evaluation and perceive certain clinical impacts such as an improvement in triaging the need for biopsy.

## Data Availability

The datasets presented in this manuscript are not readily available due to protection of patient privacy.

## Acknowledgment

The authors would like to thank Shawn Chen of the UCLA Computational Diagnostics Lab for his help in data collection.

## Notes

Manuscript received March 2023. This work was supported by the National Institute of Biomedical Imaging and Bioengineering of the National Institutes of Health under award number R21EB030691.

### Competing Interest Statement

The authors have declared no competing interest.

### Funding Statement

This work was supported by the National Institute of Biomedical Imaging and Bioengineering of the National Institutes of Health under award number R21EB030691.

### Author Declarations

IRB of University of California, Los Angeles gave ethical approval for this work.

### Summary of Updates

Introduction updated to clarify main contributions of paper. Some sections were restructured for further clarity. Paper was reformatted overall for a new journal submission.

